# Stage-dependent biomarker changes in spinocerebellar ataxia type 3

**DOI:** 10.1101/2023.04.21.23287817

**Authors:** Jennifer Faber, Moritz Berger, Wilke Carlo, Jeannette Hübener-Schmid, Tamara Schaprian, Magda M Santana, Marcus Grobe-Einsler, Dement Onder, Berkan Koyak, Paola Giunti, Hector Garcia-Moreno, Cristina Gonzalez-Robles, Manuela Lima, Mafalda Raposo, Ana Rosa Vieira Melo, Luís Pereira de Almeida, Patrick Silva, Maria M Pinto, Bart P. van de Warrenburg, Judith van Gaalen, Jeroen Jeroen de Vries, Gulin Oz, James M. Joers, Matthis Synofzik, Ludger Schöls, Olaf Riess, Jon Infante, Leire Manrique, Dagmar Timmann, Andreas Thieme, Heike Jacobi, Kathrin Reetz, Imis Dogan, Chiadikaobi Onyike, Michal Povazan, Jeremy Schmahmann, Eva-Maria Ratai, Matthias Schmid, Thomas Klockgether

## Abstract

Spinocerebellar ataxia type 3/Machado–Joseph disease (SCA3) is the most common autosomal dominant ataxia. In view of the development of targeted therapies for SCA3, precise knowledge of stage-dependent fluid and MRI biomarker changes is needed.

We analyzed cross-sectional data of 292 SCA3 mutation carriers including 57 pre-ataxic individuals, and 108 healthy controls from the European Spinocerebellar ataxia type 3/Machado-Joseph Disease Initiative (ESMI) cohort. Blood concentrations of mutant ATXN3 and neurofilament light (NfL) were determined, and volumes of pons, cerebellar white matter (CWM) and cerebellar grey matter (CGM) were measured on MRI.

Mutant ATXN3 concentrations were high before and after ataxia onset, while NfL continuously increased and deviated from normal 11.9 years before onset. Pons and CWM volumes decreased, but the deviation from normal was only 2.0 years (pons) and 0.3 years (CWM) before ataxia onset. We propose a staging model of SCA3 that includes an initial asymptomatic carrier stage followed by the biomarker stage defined by absence of ataxia, but a significant rise of NfL. The biomarker stage leads into the ataxia stage, defined by manifest ataxia.

The present analysis provides a robust framework for further studies aiming at elaboration and differentiation of the staging model of SCA3.

## Introduction

Spinocerebellar ataxia type 3/Machado–Joseph disease (SCA3) is the most common autosomal dominantly inherited ataxia disease worldwide. It is caused by unstable expansions of polyglutamine encoding CAG repeats in the *ATXN3* gene, resulting in the formation of an abnormally elongated disease protein.^1^ Several fluid and imaging biomarker candidates, that showed alterations before the clinical onset of ataxia have been identified in SCA3.^2-7^ After onset, ataxia in SCA3 takes a progressive course over an average of 20 years leading to increasing disability and premature death.^8^

Targeted therapies for SCA3 are being developed, and a first safety trial of an antisense oligonucleotide (ASOs) inducing cleavage of the RNA encoding *ATXN3* has been initiated in patients with SCA3 (https://clinicaltrials.gov/ct2/show/NCT05160558). In the future, preventive trials including pre-ataxic SCA3 mutation carriers will be a realistic option.^9^ For the design of such trials, a thorough understanding of the dynamics of the various biomarkers that reflect the cascade of pathological events associated with SCA3 is a crucial prerequisite.

We analysed a large cross-sectional dataset of SCA3 mutation carriers from the European Spinocerebellar ataxia type 3/Machado-Joseph Disease Initiative (ESMI) cohort that covers a long period of the disease course ranging from early pre-ataxic to late advanced phases. Our aim was to delineate fluid and MRI biomarker changes in relation to ataxia manifestation throughout the disease course. We concentrated on blood levels of ATXN3 and neurofilament light (NfL), as well as MRI-derived brainstem and cerebellar volumes. These markers reflect key pathological changes of SCA3 and are known to be abnormal before onset of ataxia.^4-7,10-12^ To assess the presence and severity of ataxia, we used the Scale for the Assessment and Rating of Ataxia (SARA).^13^ Our analysis allowed to propose a staging model of SCA3 based on changes of SARA and the biomarkers under study.

## Materials and methods

### Study participants

This prospective, longitudinal, observational cohort study is carried out at 14 sites in five European countries (Germany, Netherlands, Portugal, Spain, and United Kingdom) and the United States. Participants of the ESMI cohort undergo annual standardized assessment including a clinical examination and biosampling. MRI is performed at 11 sites. SCA3 mutation carriers, their first-degree relatives and healthy controls are eligible for inclusion.

For this analysis, we used cross-sectional data of 292 SCA3 mutation carriers and 108 healthy controls of whom at least one fluid or MRI biomarker result was available at Jan 31, 2022. The ESMI consortium previously published individual biomarker data separately. The present analysis is largely based on these data.^4-6^

The study was approved by the local ethics committees. Written informed consent according to the declaration of Helsinki was obtained from all participants.

### Assessments

We used the Scale for the Assessment and Rating of Ataxia (SARA)^13^ to assess the presence and severity of ataxia. Manifest ataxia was defined by a score of ≥ 3, the term “pre-ataxic” is used for all SCA3 mutation carriers with a SARA < 3. The cut-off value of three was defined by the mean plus 2 standard deviations in the initial validation study.^14^

Using a single molecule counting immunoassay, we measured plasma concentrations of expanded ATXN3.^6^ Serum concentrations of NfL were determined with an ultra-sensitive single-molecule array (Simoa) assay.^4^ One single outlier with a value of NfL 4-fold higher than all other participants was excluded.

T1-weighted MRIs were acquired using a magnetization prepared rapid gradient-echo sequence (MPRAGE, TR = 2500 ms, TE = 4.37 ms, TI = 1100 ms, flip angle = 7 deg, FOV 256 mm × 256 mm, 192 slices with a voxel size of 1 mm isotropic) on Siemens 3T scanners (Siemens Medical Systems, Erlangen, Germany). Volumes of the pons^15^, cerebellar white matter (CWM) and cerebellar grey matter (CGM)^12^ were measured and normalized by each participants total intracranial volume.

Repeat lengths of the expanded and normal alleles were determined at the Institute for Medical Genetics and Applied Genomics of the University of Tübingen. DNA samples were available in 243 of the 292 study participants. For 43 participants, information about repeat lengths was taken from medical records. In six participants, no information about repeat lengths was available.

Age of onset was defined as the reported first occurrence of gait disturbances. For SCA3 mutation carriers, not yet experiencing gait disturbances, the age of onset was calculated on the basis of CAG repeat length and the actual age^16^. The reported age of onset was missing in 15 SCA3 mutation carriers with gait disturbances. For such cases, we calculated the age of onset based on the CAG repeat length.^16^

### Statistical analysis

Statistical analysis was carried out using R version 4.1.1 (R Core Team 2022: R: A Language and Environment for Statistical Computing, R Foundation for Statistical Computing, Vienna, Austria). Descriptive analysis included the calculation of mean and standard deviation for continuous variables and frequencies (counts and percentages) for categorical variables.

Additive Gaussian regression models were used to relate fluid and MRI biomarker results in SCA3 mutation carriers to the time from ataxia onset. The functions in time were specified as a cubic P-splines with a second order difference penalty. For this, NfL concentrations and MRI volumes were z-transformed with respect to age as described before.^5^ Since SARA scores and ATXN3 concentrations in healthy controls are close to 0, no z-transformation was performed for these values, and the raw values were used.

Based on these regression models we defined the carrier, biomarker and ataxia stage. For a detailed description we refer to the results section. Fluid and imaging biomarker values, namely ATXN3, NfL and volumes of the pons, cerebellar white matter (CWM) and grey matter (CGM), as well as SARA sum scores in SCA3 mutation carriers were compared between the three stages using one-way ANOVA followed by pairwise comparisons using Tukey’s test, respectively.

The association between ataxia severity (assessed with SARA, dependent variable) and age, sex, CAG repeat length of the expanded allele, NfL levels, and pons, CWM and CGM volume as independent variables was investigated using a penalized linear regression model with LASSO penalty. Mutant ATXN3 was not included in the model, since it did not show marked temporal dynamics and would have substantially limited the number of cases. The optimal penalty parameter was determined by repeated ten-fold cross-validation (100 replications). A Box-Cox transformation with parameter λ= 0.25 was applied to the SARA sum score to approach normality.

### Data availability statement

Due to the sensitive nature of the data on rare diseases, access to the data can only be granted upon reasonable request, subject to the General Data Protection Regulation (GDPR) and any other relevant data protection laws. Please contact Jennifer Faber and Thomas Klockgether to submit a data access request.

## Results

Table 1 shows demographic and genetic data. MRI results, ATXN3 concentrations, and NfL levels were available in 161, 134, 327 participants, respectively, with an overlap between all three markers in 38 participants, between ATXN3 and MRI in 39, between NfL and MRI in 96, and between ATXN3 and NfL in 125 participants.

**Table 1.**
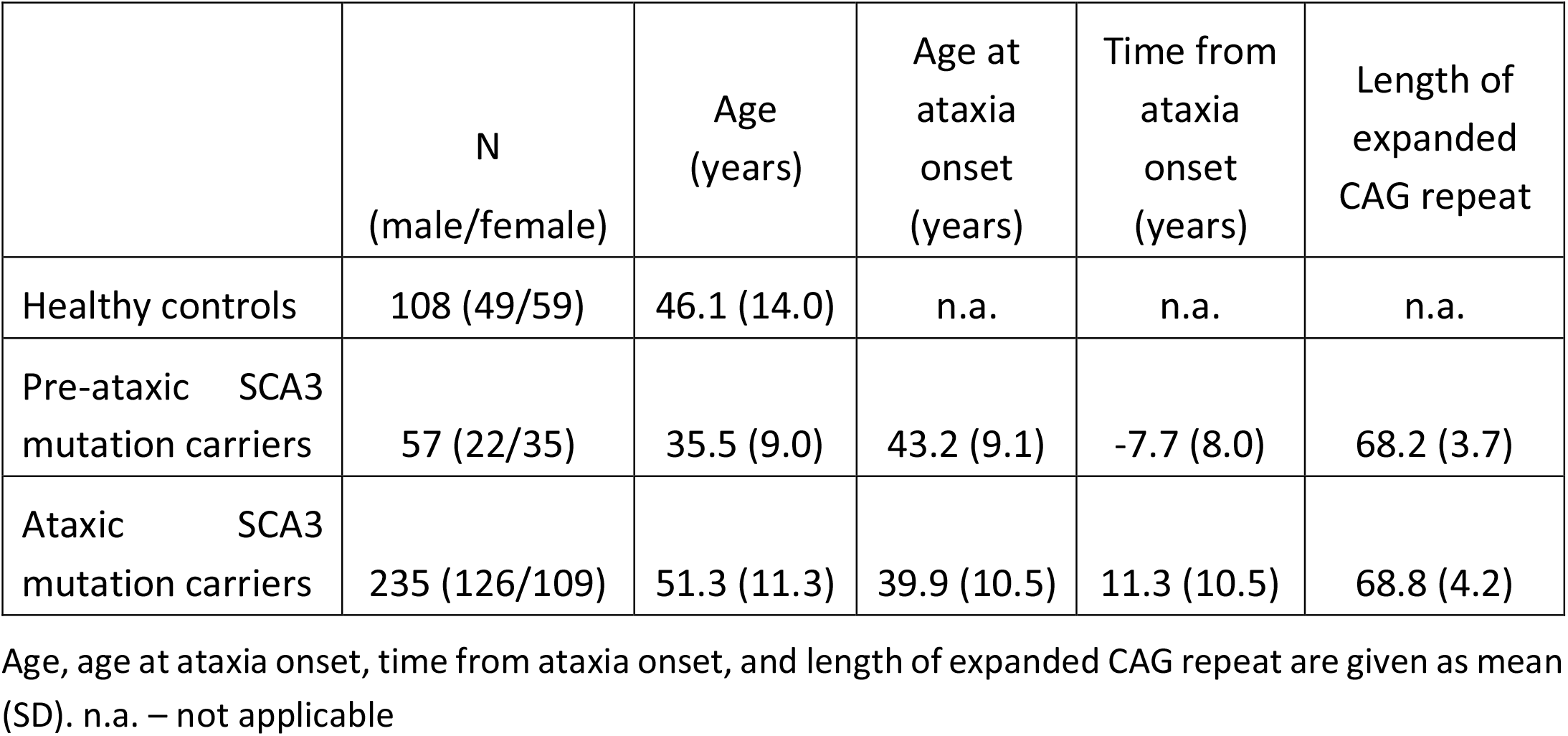
Demographic and genetic data of study participants.

|  | N<br>(male/female) | Age<br>(years) | Age at<br>ataxia<br>onset<br>(years) | Time from<br>ataxia<br>onset<br>(years) | Length of<br>expanded<br>CAG repeat |
| --- | --- | --- | --- | --- | --- |
| Healthy controls | 108 (49/59) | 46.1 (14.0) | n.a. | n.a. | n.a. |
| Pre-ataxic SCA3<br>mutation carriers | 57 (22/35) | 35.5 (9.0) | 43.2 (9.1) | -7.7 (8.0) | 68.2 (3.7) |
| Ataxic SCA3<br>mutation carriers | 235 (126/109) | 51.3 (11.3) | 39.9 (10.5) | 11.3 (10.5) | 68.8 (4.2) |

Age, age at ataxia onset, time from ataxia onset, and length of expanded CAG repeat are given as mean (SD). n.a. – not applicable

Changes of SARA scores, fluid biomarker levels, and MRI volumes of SCA3 mutation carriers in relation to the time from ataxia onset are shown in Figure 1. SARA scores were below 3 until the onset of ataxia and increased in a sigmoidal shape thereafter. Mutant ATXN3 concentrations were high with wide variation throughout the disease course, while NfL continuously increased during the pre-ataxic period and reached a plateau after ataxia onset. Overlap of the NfL 95% CIs of SCA3 mutation carriers with the interval of mean ± 2 SD of controls ended 11.9 years before onset. Pons and CWM volumes also started to decrease before ataxia onset, but the overlap of the 95% CI of SCA3 mutation carriers and the interval of mean ± 2 SD in controls ended only 2.0 years (pons) and 0.3 years (CWM) before ataxia onset. CGM volume only slightly decreased and stayed within the ± 2 SD range around the mean of controls during the entire disease course.

**Figure 1:**
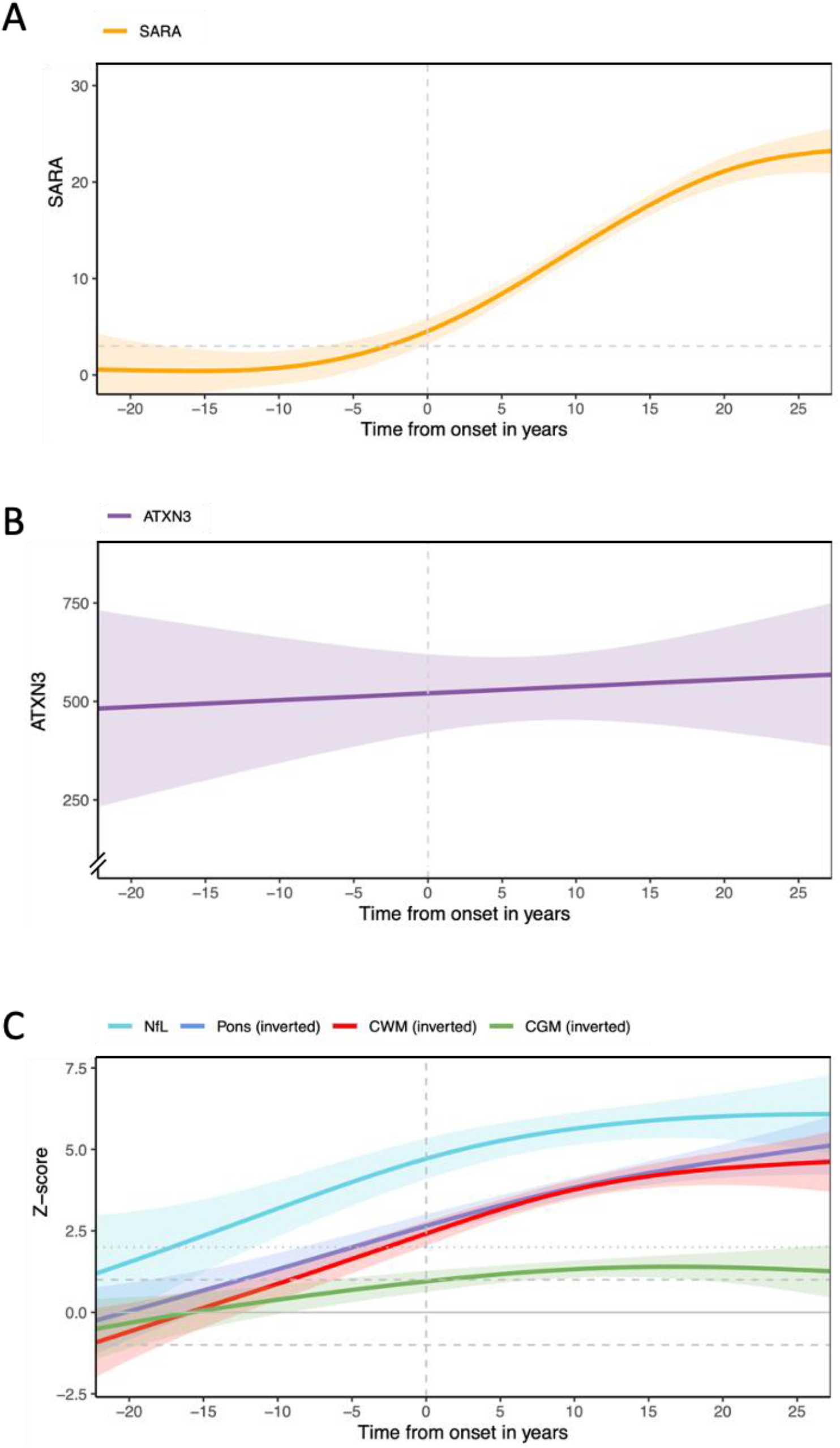
Scale for the Assessment and Rating of Ataxia (SARA) scores, fluid and MRI biomarker data in SCA3 mutation carriers in relation to time of ataxia onset. Data were analyzed with additive Gaussian regression on a time scale defined by ataxia onset. The time of ataxia onset is indicated with a vertical dashed line in all graphs. The estimated 95% CIs are shown by the shaded areas around the curves. (A) SARA sum score. The SARA cut-off of 3 defining manifest ataxia is given as a dashed horizontal line. (B) Plasma concentrations of elongated ATXN3. Data are given in ng/ml. (C) Serum concentrations of neurofilament light (NfL), MRI volumes of the pons, cerebellar with matter (CWM) and grey matter (CGM). Data were z-transformed in relation to healthy controls of same age. Y-axis of volume values is inverted for better comparability of volume loss and NfL increase. Mean of healthy controls is given as a horizontal line, the 1 SD range by dashed, and the 2 SD range by dotted lines.

Based on the temporal sequence of biomarker changes, we defined the following disease stages: (i) The carrier stage includes pre-ataxic mutation carriers without significant biomarker abnormalities other than the presence of mutant ATXN3. It is defined by SARA < 3 and an NfL z-score < 2. We chose NfL as a criterion, as the preceding analysis showed that levels of NfL were the first of the studied biomarkers to rise. (ii) The biomarker stage includes pre-ataxic mutation carriers with significant biomarker changes. It is defined by SARA < 3 and an NfL z-score ≥ 2. (iii) The ataxia stage includes ataxic mutation carriers. It is defined by SARA ≥ 3 (Figure 2).

**Figure 2:**
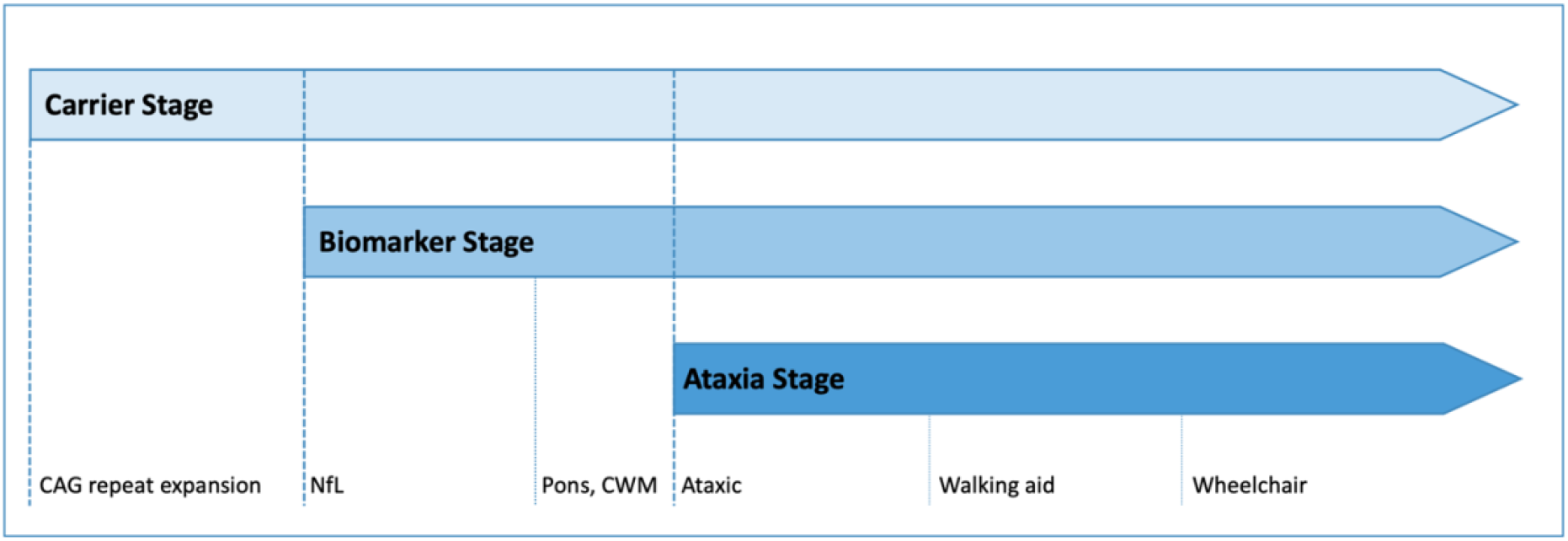
Staging model of SCA3. Proposed staging model of SCA3 based on the studied fluid and MRI biomarker data. The model includes an initial asymptomatic carrier stage followed by the biomarker stage defined by absence of ataxia, but a significant rise of NfL. The biomarker stage leads into the ataxia stage, which is defined by manifest ataxia. Following previous suggestions, the ataxia stage is further subdivided into three substages defined by milestones of gait deterioration.^30^

In the carrier stage, z-scores of MRI volumes of all SCA3 mutation carriers were > -2. In the biomarker stage, pons and CWM volume z-scores in 2 out of 11 (18 %) and CGM volume z-scores in 3 out of 11 (27 %) mutation carriers were ≤ -2. In the ataxia stage, NfL z-scores were ≥ 2 in 174 of 190 (92 %) mutation carries. Further, pons volume z-scores in 77 of 86 (90 %), CWM volume z-scores in 78 of 86 (91 %), and CGM volume z-scores in 26 of 86 (30 %) were ≤ -2.

Changes of SARA and the analyzed biomarkers in each stage are shown in Figure 3. Levels of mutant ATXN3 did not differ between the carrier, biomarker and ataxia stage in SCA3 mutation carriers. SCA3 mutation carriers in the biomarker stage showed by definition significantly increased NfL z-scores compared to the carrier stage, while SARA as well as pons, CWM and CGM volumes did not differ between the carrier and biomarker stages. SARA and all biomarkers except ATXN3 differed between the ataxia and carrier stage.

**Figure 3.**
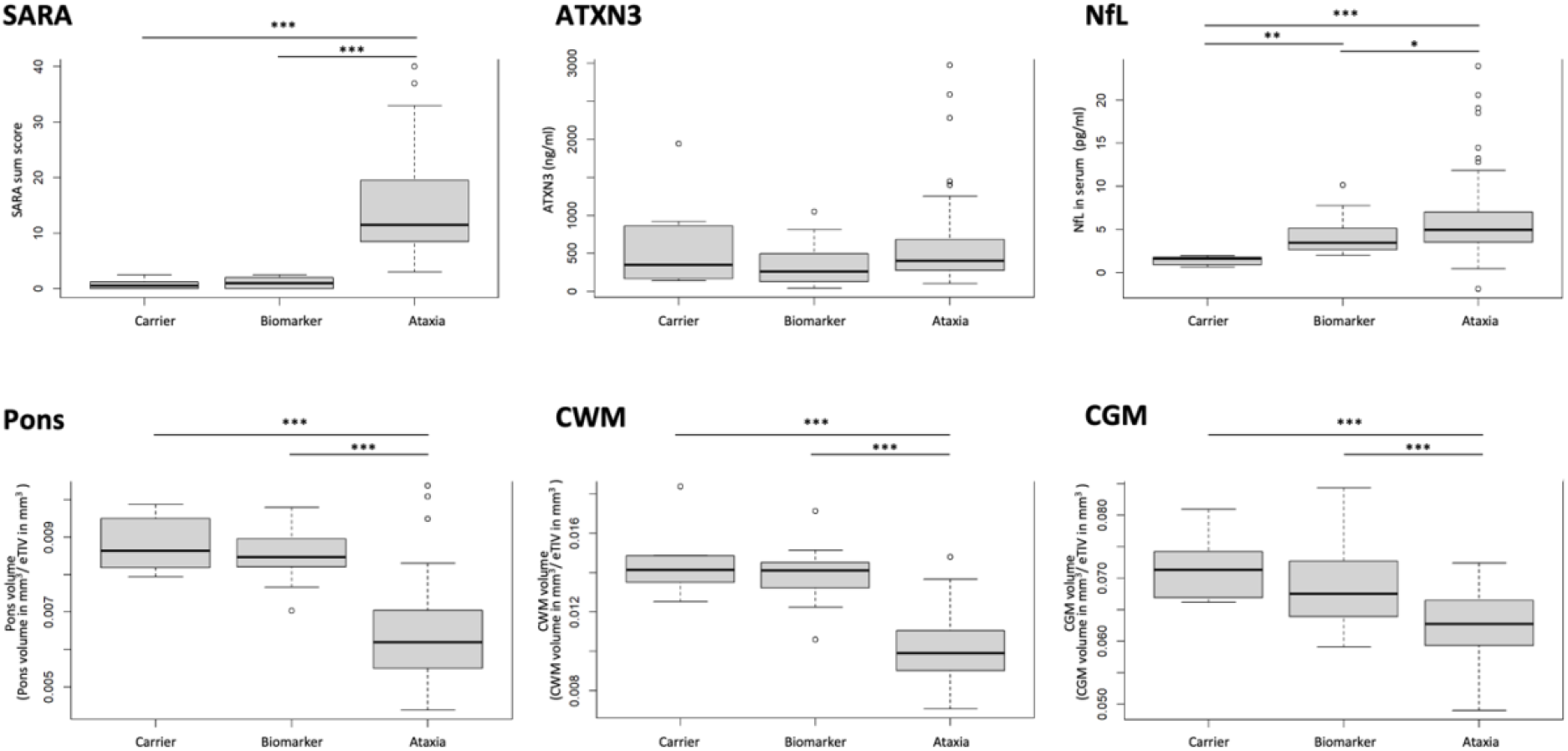
Scale for the Assessment and Rating of Ataxia (SARA) scores, fluid and MRI biomarker data in the carrier, biomarker and ataxia stage of SCA3. Data were analyzed with one-way ANOVA followed by pairwise comparisons using Tukey’s test * p<0.01; **p<0.001. NfL - neurofilament light, CWM – cerebellar white matter, CGM – cerebellar grey matter, eTIV – estimated intracranial volume.

A regression model including age, sex, CAG repeat length of the expanded allele, NfL, and MRI volumes explained 73.9% of the variability of ataxia severity, as measured by SARA. The order of selection into the penalized model, reflecting the contribution to the model from high to low, was CWM volume, age, pons volume, NfL, CAG repeat length, and CGM. A model that did not include NfL and MRI volumes explained only 60.4% of SARA.

## Discussion

Using cross-sectional data from 292 SCA3 mutation carriers from the ESMI cohort, we estimated the temporal order and extent of fluid biomarker and MRI volume changes along the disease course of SCA3. Based on the observed biomarker changes and manifestation of ataxia, we drafted a staging model of SCA3 that includes an initial asymptomatic carrier stage followed by the biomarker stage defined by absence of ataxia, but a significant rise of NfL. The biomarker stage leads into the ataxia stage, which is defined by manifest ataxia.

The present analysis is limited by its cross-sectional design. Therefore, supplementation by longitudinal data is needed. However, even with long-term studies only short sections of the entire disease course that starts before ataxia onset and spans over approximately 40 years can be covered. Although ESMI is one of the largest SCA3 cohorts worldwide, the amount of biomarker data available for analysis was limited. To improve statistical power in future analyses, merging of ESMI data with data from other cohorts is mandatory.

Plasma concentrations of the elongated disease protein ATXN3 were high detectable throughout the entire disease course and did not differ between the different stages. This is best explained by the fact that elevation of mutant ATXN3 levels is the direct consequence of the gene mutation causing SCA3. Nature and extent of the changes characterize mutant ATXN3 as a trait rather than a progression biomarker. We therefore did not use ATXN3 as a criterion for the definition of disease stages.

The rise of NfL preceded ataxia onset by 11.9 years. This agrees with previous reports on NfL data of ESMI participants,^4,7^ as well as findings in other cohorts.^17,18^ As the rise of NfL marks the first currently detectable damage to the nervous system in SCA3, we used NfL as a criterion to define the biomarker stage in pre-ataxic individuals.

While NfL is supposed to reflect the rate of degeneration,^4,19,20^ MRI regional volume loss rather represents the cumulated result of degeneration explaining why volume loss followed the rise of NfL, and why MRI volumes did not differ between the carrier and biomarker stage on a group level.^17^ Nevertheless, pons and CWM volumes showed a continuous decrease and deviated from normal 2.0 and 0.3 years before ataxia onset. Consequently, they may be considered for the identification of mutation carriers close to the clinical onset. CGM volume loss overall was less pronounced and most prominent in the ataxia stage. These observations are in line with autopsy findings that show, unlike most other SCAs, relative sparing of cerebellar cortex in SCA3.^21^ The strong involvement of the CWM volume corroborates previous findings of prominent white matter loss in patients with SCA3,^22,23^ and is in line with reports of early oligodendrocyte pathology in mouse models of SCA3.^24,25^ Pons volume showed an almost linear decline along the entire disease range, denoting it as a potential marker of disease progression.

A previous study in SCA3 patients showed that ataxia severity, as measured by SARA, can be predicted by CAG repeat length and age.^26^ The present data show that the accuracy of the prediction was substantially improved by adding NfL levels as well as pons and CWM volumes to the prediction model underlining the biological relevance of these markers.

The present data allowed for the first time to draft of a data-driven model of disease stages for SCA3. While the disease was previously divided into a pre-ataxic and ataxic stage, we propose a more differentiated model similar to that recently presented for Huntington’s disease (HD).^27^ We defined the onset of biomarker stage by the rise of NfL. However, it is possible that changes of other biomarkers indicating incipient damage to the nervous system precede the rise of NfL. Further studies including additional fluid and imaging biomarker data, such as MR spectroscopy and diffusion imaging,^11^ may allow to further subdivide the biomarker stage. In the present model, we have not introduced a clinical sign or symptom stage preceding the final clinical stage like in the HD model.^27^ Yet, this might be considered reasonable, since such signs and symptoms, like oculomotor dysfunction, have been described in pre-ataxic SCA3 mutation carriers.^14,28,29^ The ataxia stage has previously been subdivided into three stages defined by milestones of gait deterioration leading to further differentiation of the model.^30^

The present staging model of SCA3 is to be considered as first proposal that needs to be further refined and extended based on more data and broad consensus. Nevertheless, it provides a robust framework for further studies aiming at elaboration and differentiation of the model.

## Acknowledgements

TK, MS, LS, JI, BvdW are members of the European Reference Network for Rare Neurological Diseases (ERN-RD, project number 739510). The ESMI consortium acknowledges Ruth Hossinger for the project management of the ESMI project and for all contributions made towards the success of this project.

